# Evolution of COVID-19 pandemic: Power-law growth and saturation

**DOI:** 10.1101/2020.05.05.20091389

**Authors:** Soumyadeep Chatterjee, Ali Asad, B. Shayak, Shashwat Bhattacharya, Shadab Alam, Mahendra K. Verma

## Abstract

In this paper, we analyze the real-time infection data of COVID-19 epidemic for 21 nations up to June 30, 2020. For most of these nations, the total number of infected individuals exhibits a succession of exponential growth and power-law growth before the flattening of the curve. In particular, we find a universal 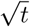 growth before they reach saturation. However, at present, India, which has *I*(*t*) ~ *t*^2^, and Russia and Brazil, which have *I*(*t*) ~ *t*, are yet to flatten their curves. Thus, the polynomials of the *I*(*t*) curves provide valuable information on the stage of the epidemic evolution, thus on the life cycle of COVID-19 pandemic. Besides these detailed analyses, we compare the predictions of an extended SEIR model and a delay differential equation-based model with the reported infection data and observed good agreement among them, including the 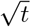 behaviour. We argue that the power laws in the epidemic curves may be due to lockdowns.

## 1 Introduction

As of July 05, 2020, COVID-19 pandemic has infected more than 11.4 million of the human population and caused 0.34 million deaths. The world economy is in tatters. Therefore, understanding the progression of the pandemic is extremely crucial. In the present paper, we analyze the publicly-available national COVID-19 infection data [40] up to June 30, 2020. We observe that the COVID-19 infection curves for many nations exhibit power-law growth after exponential growth. We compare the reported data with model predictions and observe a good agreement among them.

To understand and forecast epidemics, epidemiologists have made many models [16, 4, 11]. One of the first models is called the SIR model, where the variables *S* and *I* describe the numbers of susceptible and infected individuals, respectively. The third variable *R* represents the removed individuals who have either recovered or died. An advanced model, called SEIR model, includes *exposed* individuals, *E*, who are infected but not yet infectious [4, 11].

SARS-CoV-2 is one of the seven human coronaviruses which have been identified so far. It is the most dangerous among all of these because of its highly infectious nature and its lethality. *Asymptomatic carriers*, individuals who do not exhibit any symptoms, have carried the virus to far off places where it has spread rapidly [20]. Even symptomatic patients manifest symptoms two to three days after turning transmissible. To stop the spread of the deadly virus, various nations have employed lockdowns, mandatory social distancing, quarantines for the affected, etc.

Despite the difficulties stated above, many models are able to describe the COVID-19 pandemic data quite well. Peng et al. [28] constructed a generalized SEIR model with seven-variables (including quarantined and death variables) for the epidemic spread in China. Their predictions are in good agreement with the present data. López and Rodo [21] formulated an extended version of this model to analyse the spread of the pandemic in Spain and Italy. Earlier, Cheynet [8, 9] had developed a code to simulate this model. Hellewell et al. [14] studied the effects of isolation on controlling the COVID-19 epidemic. Chinazzi et al. [10] analyzed the effects of travel restrictions on the spread of COVID-19 in China and in the world using the global metapopulation disease transmission model. Mandal et al. [24] constructed model for devising intervention strategies in India. Shayak et al. [35] have constructed delay differential equation (DDE) model for the spread of COVID-19; this model takes into account the pre-symptomatic period and predicts a path to the end of the epidemic.

Due to the above complex issues in the epidemic models of COVID-19, many researchers have consciously focussed on the data and attempted to extract useful information from them. It has been observed that the analysis of the pandemic provides important clues that may be useful for its forecast. In particular, Ziff and Ziff [42], Komarova and Wodarz [17], Manchein et al. [23], Blasius [5], Marsland and Mehta [26], Li et al. [19], Singer [36], Beare and Toda [2], and Cherednik and Hill [7] analyzed the reported count of total infections (*I*(*t*)) in various nations and observed power-law growth after the exponential regime. Verma et al. [37] analyzed the data of 9 nations up to May 4, 2020 and showed that the *I*(*t*) goes through power laws, *t*^3^, *t^2^, t* and 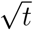, in temporal sequence before flattening out. They observed that by April 7, 2020, most of the nations studied by them were either still in the exponential regime or the power-law regime, except China and South Korea, which had flattened their curves. However, by June 30, many nations had flattened their epidemic curves. Unfortunately, some of them appear to have a second wave of infections. Besides above results, Prakash et al. [32] reported a linear growth of *I*(*t*) after early exponential growth.

Schüttler et al. [34] analyzed the daily death counts for various nations and observed that their probability distributions appear to follow a Gaussian profile. Marsland and Mehta [26] observed that the error function provides best fit to the total count, *I*(*t*); this observation follows from Schüttler et al. [34]’s analysis. There are epidemic growth models based on population growth [11, 41]. COVID-19 spread via asymptomatic carriers leads to a network formation. Hence, network-based epidemic growth models may be useful for modelling COVID-19 pandemic. Marathe and Vullikanti [25] review computational epidemiology with a focus on epidemic spread over a network.

In this paper, we analyze the COVID-19 infection data up to June 30, 2020 for 21 nations and observe that all the nations are following transition from exponential to power-law growth in infection counts. Most of the 21 nations have flattened their curves with several exceptions (for example, Russia). We also show that three epidemics—Ebola, COVID-19, MERS— have similar evolution: exponential growth, power-law growth, and then flattening of the curve. In addition, we compare the predictions of an extended SEIR model [21] and a delay-differential equation model [35] with the real-time data and observed good agreement among them. Finally, we present a summerized overview of the universal temporal variations of the COVID-19’s epidemic curve.

The structure of the paper is as follows: in Sec. 2 we analyse the COVID-19 data for 21 leading nations and observe power law growth for them after the exponential growth. The evolutions of Ebola, MERS, and COVID-19 are compared in Sec. 3. The predictions of two models of COVID-19 pandemic are compared with the observed data in Sections 4 and 5. In Sec. 6 we present and explain a general viewpoint depicting the life cycle of COVID-19 pandemic. We conclude in Sec. 7.

## 2 Data analysis of COVID-19 epidemic

In this section, we present our results based on a comprehensive data analysis of COVID-19 cases for 21 countries (see Table 1) up to June 30, 2020. The majority of the countries in our analysis include those with a large number of COVID-19 cases, including USA, Italy, Germany, China, and India. For a complete study, we also include countries with a relatively smaller number of cases such as Sri Lanka and Hong Kong. We used the real-time data available at *worldOmeter* [40] and chose the starting date (see Table 1) as the one from which the number of infected cases increased rapidly. Corona Resource Center [15] too is an important repository for COVID-19 data.

**Table 1:**
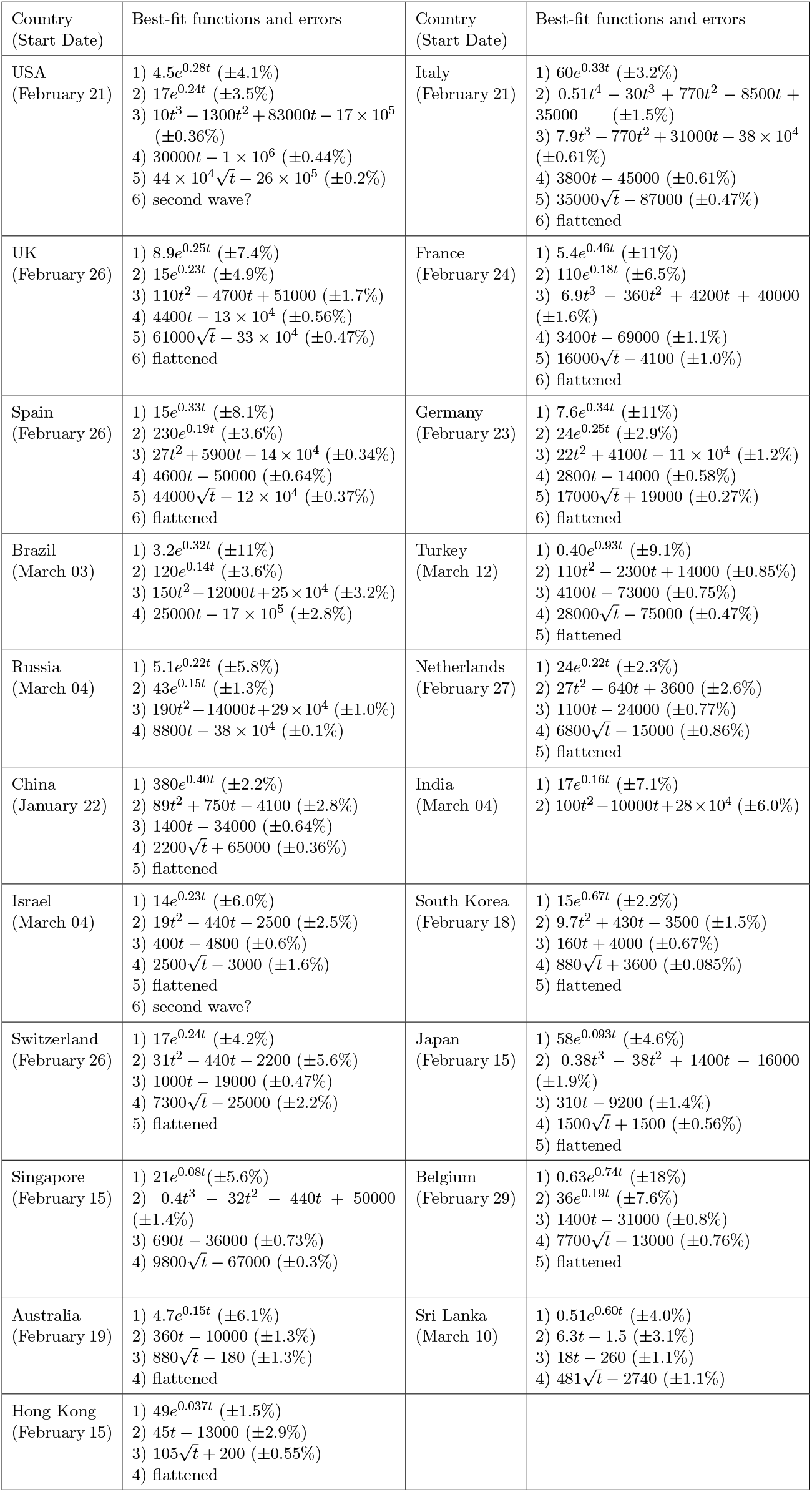
The best-fit curves along with their relative errors for the COVID-19 data for 21 countries.. The best-fit curves are shown in Figs. 1. The start date for the plots of Fig. 1 are listed below the names of the countries.

We analyze the evolution of cumulative number of infected cases, which is denoted by *I*(*t*), with time in days, denoted by *t*. For all the *I*(*t*) curves, we compute the derivatives *İ*(*t*) using Python’s *gradient* function. These derivatives indicate the daily count of the infected cases. Note that *İ*(*t*) exhibit lower fluctuations than the measured daily counts due to smoothing. In Fig. 1, we exhibit the plots of *I*(*t*) (red curves) and *İ*(*t*) (blue curves) in *semi-logy* format for all the 21 countries.

**Figure 1:**
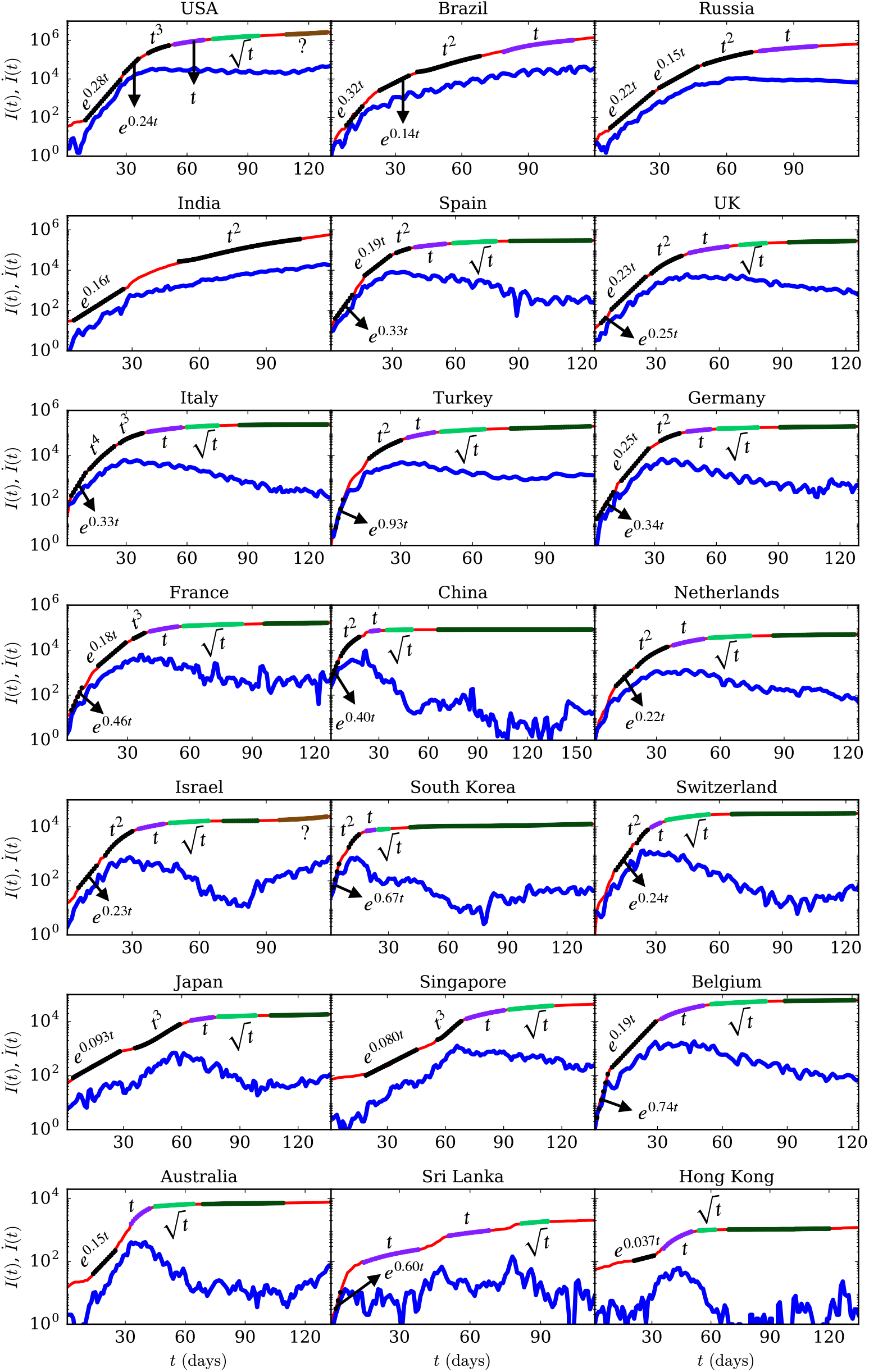
(color online) For the COVID-19 epidemic, the *semi-logy* plots of total infected individuals (*I*(*t*)) vs. time (*t*) (red curves) for 21 countries. We also plot *İ*(*t*) vs. *t* (blue curves). The dotted curves represent the best-fit curves, which are exponential or polynomial functions (see Table 1). The dark-green curves represent the flattened regions of the epidemic.

We find that a single function does not describe the *I*(*t*) curves; hence, we compute best-fit curves for different parts of *I*(*t*) by employing exponential and polynomial functions. We used Python’s *polyfit* function to compute the best-fit curves. These curves are listed in Table 1 along with the relative errors between the original data and the fitted data. However, we exhibit only the leading power laws of the polynomials in the plots of the figures.

Initially, all the countries exhibit exponential growth (*I*(*t*) = *A* exp(*βt*)), which is expected. It is worth mentioning that the *I*(*t*) plots for USA, UK, France, Spain, Germany, Russia, Belgium, and Brazil have two exponential functions for the fit. For example, the *I*(*t*) curve of UK is described by two exponential functions, ~ exp(0.25*t*) and ~ exp(0.23*t*). The quantity *β* is proportional to the growth rate. The value of *β* varies for different countries as it depends on factors such as population density, immunity level of the population, climate, local policy decisions (social distancing, lockdowns, testing capacity), etc.

In the exponential regime, the daily infection count is directly proportional to the cumulative infections, that is, *İ* ≈ *βI*. The cumulative case count doubles in time *T* = (log2)/*β* in this regime, For Italy, *β* = 0.33, resulting in *T* ≈ 2 days, which means Italy’s *I*(*t*) doubled every two days in the early phase (February 22 to March 01).

Next, the curves transition to the regimes that are best described by polynomials and can be approximated as power laws. In Fig. 1, we report the leading terms of the best-fit polynomials as power laws (also see Table 1). However, the effective exponent of power law tends to be larger than highest degree of the polynomial. For example, the power law for India is 3.5 which is higher than the highest degree of polynomial, 2. We observe that among the 21 countries considered, 10 of them—Spain, UK, Germany, France, China, Turkey, Netherlands, Israel, South Korea, Switzerland—have flattened their curves via 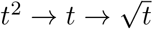. USA, Japan, Singapore exhibit transition through 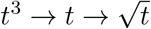, however, Australia, Sri Lanka, Hong Kong, and Belgium have reached plateau via 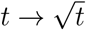. For saturation, Italy took a long path via 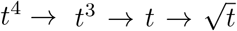. At present, Brazil and Russia have reached only the linear phase after *t*^2^ regime. Unfortunately, India is still at *t*^2^ regime. The three nations—India, Russia, Brazil—are some distance away from saturation. We also remark that USA and Israel may be getting to a second wave due to the rising daily cases in the recent times. In Fig. 1 the possible second wave is marked as brown curves. Note that the variations in the power laws and route to saturation are due to various factors: extent of lockdowns and social distancing, ease of unlocking, travel restrictions, country size, level of immunity of the population, awareness, etc. (see Sec. 6). We make a cautionary remark that the coefficients of the polynomials depend quite critically on the choice of endpoints of the fit. Our observations of power-law growth are in general agreement with earlier results [42, 17, 23, 5, 26, 19, 36, 2, 37].

At the plateau of the epidemic curve, *I*(*t*) = constant and *İ*(*t*) = 0. However, most countries claim that they have flattened their curves even when *İ*(*t*) ≠ 0, but it is small (less than 1000). Due to the various complex factors present at the moment, such fluctuations are considered to be negligible. Here, the flattening of the curve is to be taken in this spirit. In Fig. 1, the flattened regions of the epidemic curves are shown using dark-green curves.

In Fig. 1, in the exponential regime, the *İ*(*t*) curves (daily counts) are nearly parallel to the *I*(*t*) curves. It means that *İ* increases exponentially in the beginning, similar to *I*(*t*). Subsequently, the curves transition to power-law regimes. As discussed by Verma et al. [37], the power law can be approximated as *I*(*t*) ~ *At^n^* and *İ*(*t*) ~ *I*^1-1/^*^n^*, which is slower than I for the exponential regime. We also remark that for large *n*, *İ*(*t*) ~ *I*, similar to exponential function. The linear growth regime has an interesting property. In this regime, *İ* ≈ const., that is, constant daily infection count. The daily infection count starts to decrease after the linear regime; hence linear regime is the transition point.

We also analyze the data of cumulative infected individuals in the entire world. In Fig. 2, we plot *I*(*t*) and *İ*(*t*) versus time in *semi-logy* format. Note that the initial epicenter of the COVID-19 outbreak was in China, and then it shifted to Europe and then to USA. Therefore, we divide the plot in two parts. In the first part [Fig. 2(a)], we illustrate cases that belong mostly to China. After approximately thirty days of outbreak (around February 20), *I*(*t*) for China starts to saturate. In Fig. 2(b), we exhibit the 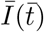 curve after *t* = 41 (March 02) when China had achieved flattening of the curve. In Fig. 2(b), 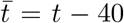 and 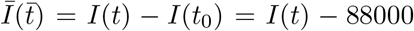 due to the coordinate shifts. Both the plots exhibit exponential and power law regimes, but 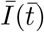 is yet to flatten (see Table 2). We hope that there is no third part to this curve.

**Figure 2:**
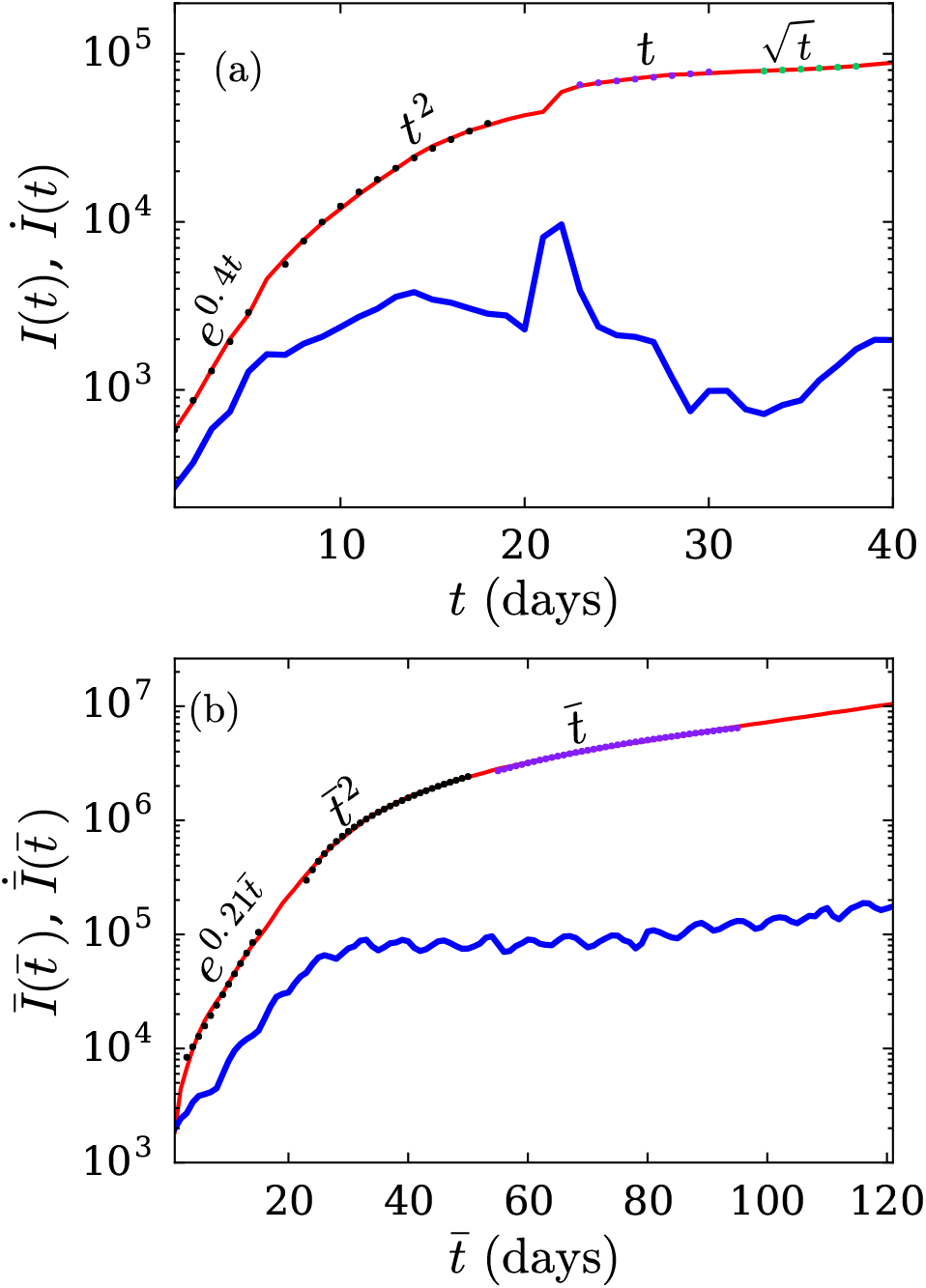
(color online) For the COVID-19 epidemic, the *semi-logy* plots of total infected individuals (*I*(*t*)) vs. time (*t*) (red curves) for (a) Part 1: from January 22 to March 01 (comprising mostly of China), (b) Part 2: from March 02 to June 30 (world other than China). In Part 2, 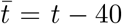 and 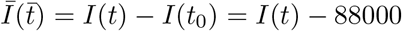. The blue curves in the plot describe the derivatives of *I*(*t*). The dotted curves represent the best-fit curves listed in Table 2.

**Table 2:** The best-fit curves along with their relative errors for the COVID-19 data for the world. The best-fit curves are shown in Fig. 2. Part 1 of the curve (Fig. 2(a)) is from January 22 to March 01, while Part 2 (Fig. 2(b)) is from March 02 to June 30. For part 2, 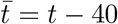 and 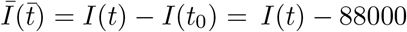, that is, time is measured from March 02.

| Part<br>(Start Date) | Best-fit functions and errors |
| --- | --- |
| Part 1<br>(January 22) | 1) $390e^{0.40t}$ ($\pm 2.3\%$)<br>2) $90t^2 + 750t - 4100$ ($\pm 2.7\%$)<br>3) $1800t + 25000$ ($\pm 0.89\%$)<br>4) $13000\sqrt{t} + 6500$ ($\pm 0.29\%$) |
| Part 2<br>(March 02) | 1) $4400e^{0.21\bar{t}}$ ($\pm 6.8\%$)<br>2) $340\bar{t}^2 + 53000\bar{t} - 11 \times 10^5$ ($\pm 1.48\%$)<br>3) $94000\bar{t} - 24 \times 10^5$ ($\pm 1.18\%$) |

The transition from the exponential to power-law behaviour is expected from the nature of the *I*(*t*) curve. The *I*(*t*) curve is convex during the exponential growth phase, that is, its center of curvature is upward. However, the curve must turn concave for it to flatten. This transformation occurs via a sequence of growth phases: power-law, linear, square-root, and then flat. The curve transitions from convex to concave in the linear regime for which the radius of curvature is infinite. In Sec. 7 we argue that the power-law behaviour is possibly due to lockdown and social distancing.

We remark that the death count due to COVID-19 also exhibits similar behaviour as the infection count *I*(*t*). It is expected because a fraction of infected individuals, unfortunately, die. However, we expect a small time delay between the death time series and the infection time series. Some researchers have attempted to fit the *I*(*t*) and death counts with error functions [26, 34].

The above analysis shows that we can track the development of the epidemic locally in time. The best-fit curves in particular segments provide the status of the epidemic. For example, if we have reached the linear regime, then we are not far from flattening the curve. Similarly, a 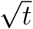 regime indicates that the flattening of the curve has begun. Thus, simple data analytics described above has significant predictive power. We expand this issue further in Sec. 6 using a schematic picture.

In the next section we compare the functional behaviour of COVID-19’s *I*(*t*) curve with those of other major epidemics.

## 3 Comparison of COVID-19 with other epidemics

A natural question is whether the epidemic evolution of COVID-19 differs from the spread of Ebola and MERS (Middle Eastern Respiratory Syndrome). In this section we perform a comparative study of Ebola, MERS, and Covid-19 epidemics. We digitized data for these epidemics for their respective time periods: MERS [30] from May 01, 2013 to April 30, 2015; Ebola [22] from May 01, 2014 to April 30, 2015; and Covid-19 [40] from January 22, 2020 to June 30, 2020. In Fig. 3, we plot *I*(*t*) vs. normalized time, *t*/*t_max_*, in a *semi-logy* format for all three epidemics. Here, *t_max_* (see Table 3) is the time span of the epidemic, except for COVID-19 for which *t_max_* is taken up to June 30.

**Figure 3:**
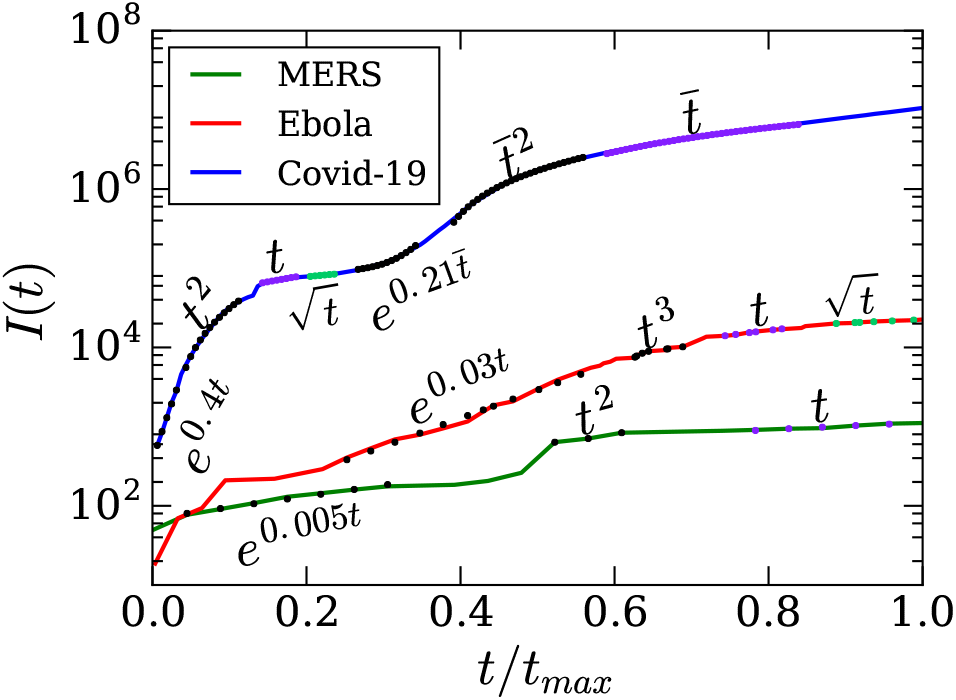
(color online) For the COVID-19, Ebola and, MERS epidemics, the *semi-logy* plots of total infected individuals (*I*(*t*)) vs. normalized time (*t*/*t_max_*). The dotted curves represent the best-fit curves using the exponential and polynomial functions (see Table 3). For COVID-19 curve, refer to Fig. 2.

**Table 3:**
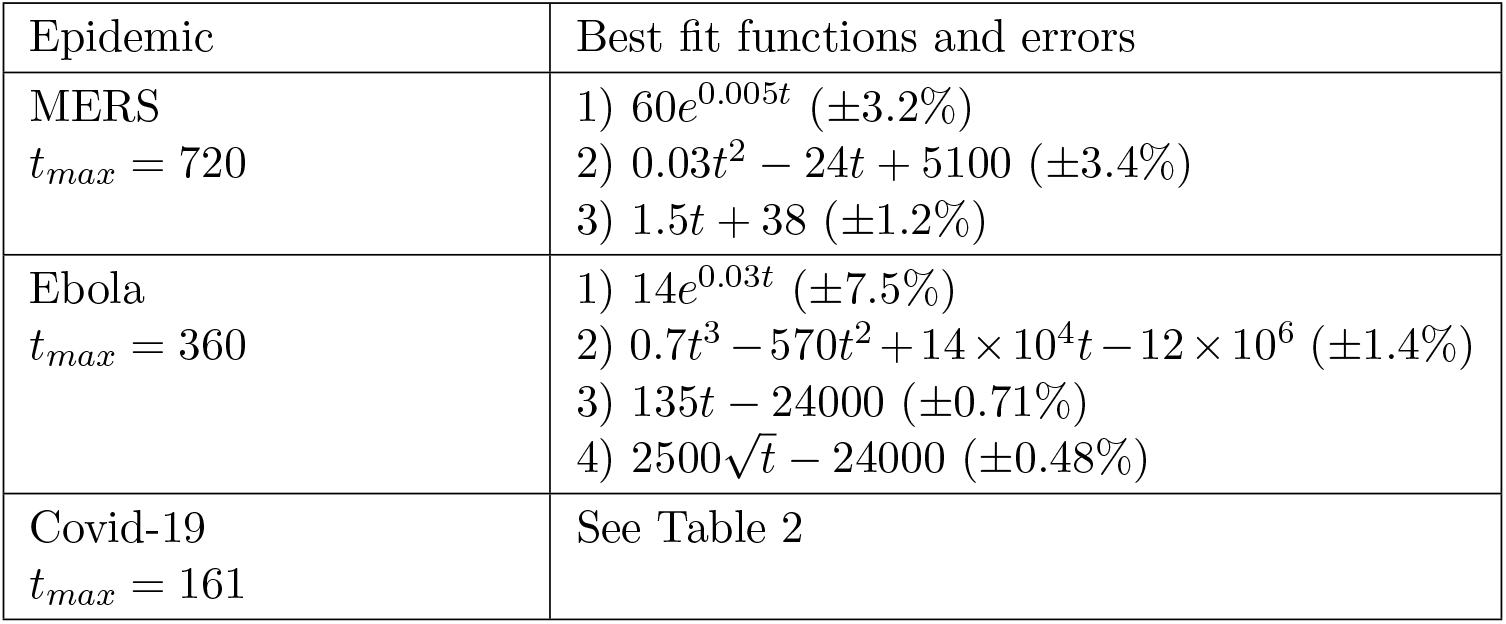
For the cumulative infections for MERS, Ebola, and COVID-19, the best-fit functions and their relative errors for various stages of evolution shown in Fig. 3.

In Fig. 3 we present the best-fit curves as dotted lines. Clearly, the three curves look similar, with regimes exhibiting exponential, power-law, and linear growth before flattening. A major difference is that COVID-19 has two subparts, which is essentially due to more spread of COVID-19 compared to other two epidemics. As shown in Table 2 and Fig. 2, the epidemic first spread in China and then in rest of the world. In contrast, the other two epidemics, Ebola and MERS, were somewhat confined.

In the next section, we present a model for COVID-19 whose predictions match for several countries.

## 4 SEIR model for COVID-19 epidemic

In Sec. 2, we analyzed the COVID-19 infection data and observed a power-law growth (followed by a linear regime near saturation) after an exponential growth. In this section, we attempt to get some insights about this transition using SEIR model [4, 11, 21, 18, 29]. Note the other important epidemic models are regression models [31, 13], ARIMA forcasting model [1, 3,12], SIR model [16, 33], etc. All these models have been frequently and successfuly used to analyse the transmission dynamics of COVID-19. For example, Labadin and Hong [18] used this model to predict the second confirmed case in Malaysia.

Recently, Peng et al. [28] constructed a generalised SEIR model for the spread of SARS-Cov-2 virus in China. López and Rodo [21] modified Peng et al. [28]’s model to analyze the data of Spain and Italy up to the end of March. In this section, we will discuss a simplified version of López and Rodo [21]’s SEIR model and fit it with the real-time data of USA, Italy, Spain and Japan till June 30 2020.

In the model, we assume the disease transmission to take place only among humans. Further, the natural birth and death rates are assumed to be negligible. We divide the total population (*N*) at a certain place at time t into seven categories: Susceptible (*S*(*t*)), Exposed (*E*(*t*)), Infected (*I*(*t*)), Recovered (*R*(*t*)), Insusceptible (*P*(*t*)), Quarantined (*Q*(*t*)) and Dead (*D*(*t*)). Here, *Q*(*t*) is the number of confirmed infected cases at time t. The evolution equations of the seven categories are:

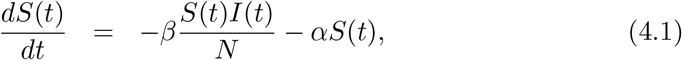

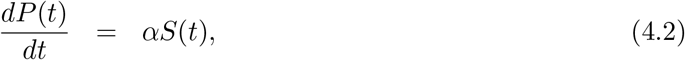

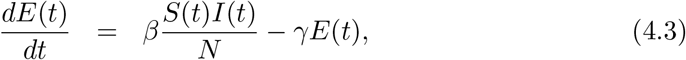

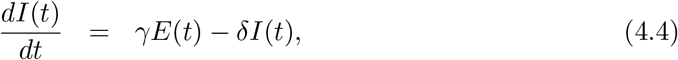

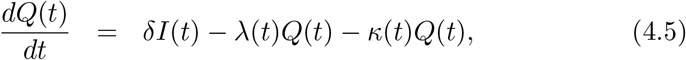

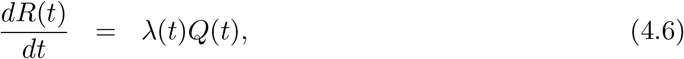

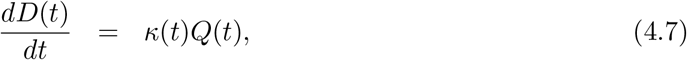

where *β*; *α*; *δ*; γ(*t*); and *κ*(*t*) are the infection, protection, average quarantine, recovery and mortality rates respectively; and γ^−1^ is the average latency period for COVID-19. The protection rate *α* is governed by the intensity of contact tracing, lockdown policies, and improvement of health facilities. The time-dependent parameters λ(*t*) and *κ*(*t*) are modeled as follows [21]:

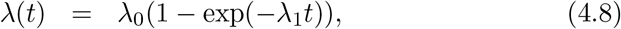

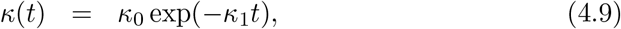

where λ_0_, *κ*_0_, λ_1_, and *κ*_1_ are constants. The functional forms in Eqs. (4.8-4.9) are chosen in such a way that the recovery rate saturates and the death rate vanishes with time. Note that the cumulative number of reported infected cases (denoted by *I*(*t*) in Sec. 2) is the sum of *Q*(*t*), *R*(*t*), and *D*(*t*). For further details, refer to Peng et al. [28], and Lopez and Rodo [21].

We compare the model predictions [Eqs. (4.1-4.9)] with the available data [40] for USA, Italy, Spain and Japan. For Spain and Japan, *t* = 0 is taken to be the starting date shown in Table 1. For USA and Italy, *t* = 0 corresponds to 29th and 24th February respectively. The end date for all the four countries is June 30. The initial values of *Q, R*, and *D* are taken to be the total active cases, recovered cases and deaths respectively at *t* = 0 for each country. The number of initial insusceptible cases (*P*(*t* = 0)) is assumed to be zero. We adjust the parameters {*α*, *β*, γ, *δ*, {λ_0_, λ_1_}, {*κ*_0_, *κ*_1_}} *E*(*t* = 0) and *I*(*t* = 0) such that the relative error between the model and actual data is minimized. Note that the initial condition satisfies the relation

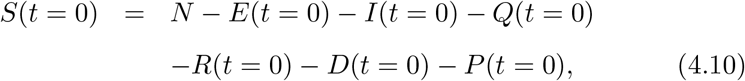

where *N* is the total population of the country.

In Fig. 4, we present the best-fit curves from the SEIR model along with the actual real-time data. In Table 4 we list the numerical values of the best-fit parameters and the relative and root mean square (RMS) errors between the predictions and data. Here, the RMS errors measure the differences between the predicted *I*(*t*) from the SEIR model and available data [40] for *I*(*t*) in logarithmic scale. Our SEIR model fits well with the data for Italy and Spain, as depicted in Fig. 4 and indicated by the low value of RMS and relative errors for these two countries. Note that for Spain and Italy, López and Rodo [21] considered natural birth and death rates in their model and obtained fits for *Q, R* and *D* seperately. In contrast, we stick to the fundamental assumption regarding natural birth and death rates of the basic SEIR model [18] and obtain the fits for *Q*(*t*) + *R*(*t*) + *D*(*t*) till June 30.

**Figure 4:**
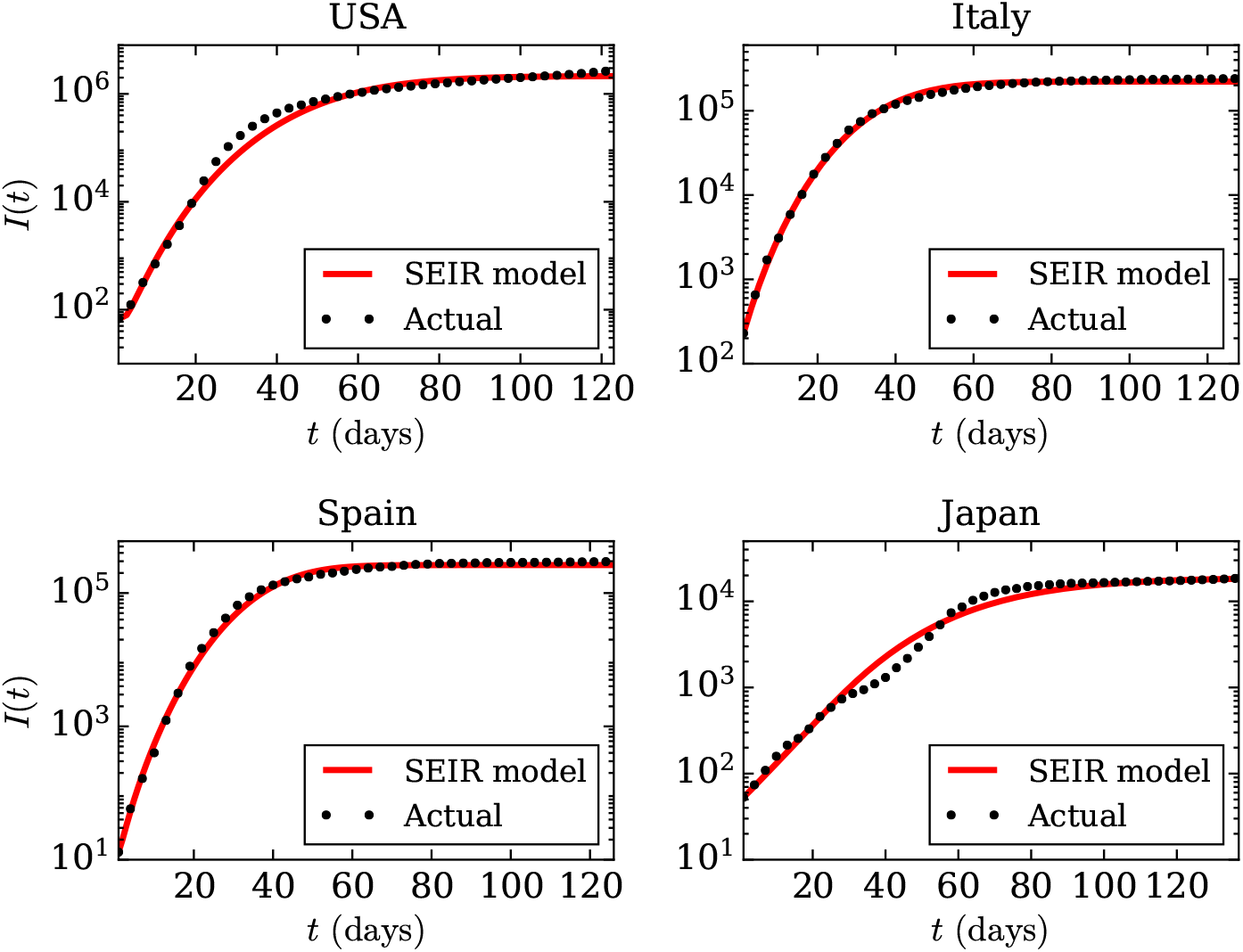
(color online) For the COVID-19 pandemic, *semi-logy* plots of total infected individuals (*I*(*t*)) vs. time (*t*) (black dotted curves) for USA, Italy, Spain and Japan from the available data at *worldOmeter* [40] till June 30. The red curves are the best-fit curves using the SEIR model described in Sec. 4.

**Figure 5:**
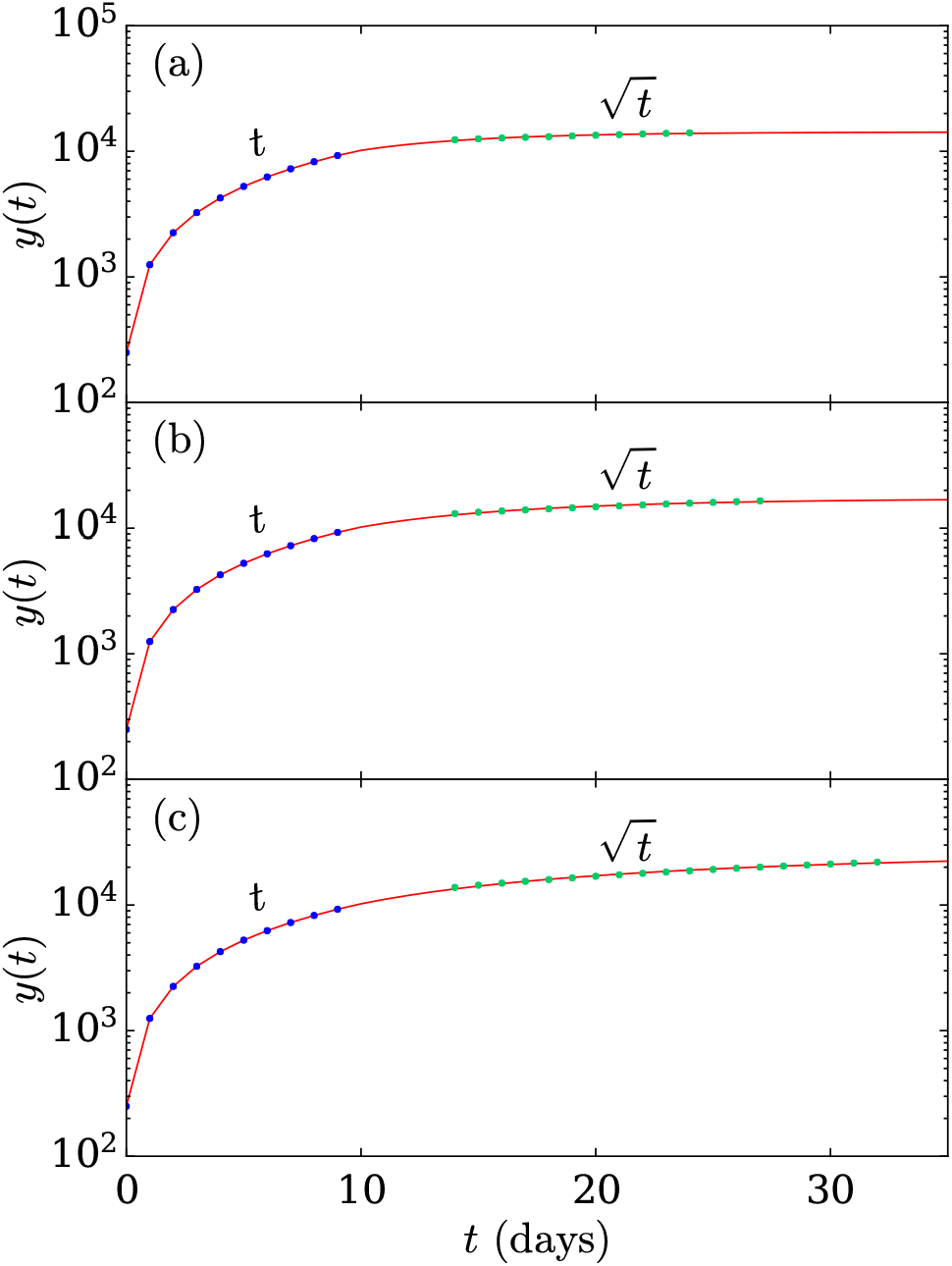
(color online) Simulation of the DDE-based model (Eq. (5.1)) with *m*_0_ being (a) 70%, (b) 80%, (c) 90% of the critical value, as explained in the Sec. 5.

**Table 4:** Best-fit numerical values of {*α*, *β*, γ, *δ*, {λ_0_, λ_1_}, {*κ*_0_, *κ*_1_}} of SEIR model for four countries with relative and root mean square (RMS) errors between the best-fit curves and actual data [40]. The best-fitt curves are shown in Fig. 4.

|  | Best-fit parameters of *SEIR* model |  |  |  |  |  |  |  | Errors |  |
| --- | --- | --- | --- | --- | --- | --- | --- | --- | --- | --- |
| Country | $\alpha$ | $\beta$ | $\gamma$ | $\delta$ | $\lambda_0$ | $\lambda_1$ | $\kappa_0$ | $\kappa_1$ | Relative (%) | RMS |
| USA | 0.027 | 7.9 | 0.11 | 1.7 | 0.8 | 0.099 | 2.0 | 2.0 | 17.1% | 0.28 |
| Italy | 0.037 | 1.4 | 0.21 | 0.42 | 0.68 | 0.045 | 0.28 | 0.045 | 5.5% | 0.063 |
| Spain | 0.016 | 1.9 | 0.87 | 0.99 | 1.3 | 0.14 | 0.25 | 0.16 | 11.2% | 0.14 |
| Japan | 0.034 | 2.1 | 0.053 | 0.25 | 0.2 | 2.2 | 0.95 | 0.2 | 16.1% | 0.21 |

Our best-fit values of parameters for Spain and Italy are nearly consistent with those of López and Rodo [21]. The model shows that high infection rates (*β*) and small average latency periods γ^−1^ try to push the cumulative number of infected cases (reported) to a large saturation value via an exponential growth. On the other hand, high protection and quarantine rates, *α* and *δ*, slow down the growth and minimize the saturation level of the cumulative infected (reported) cases. Thus, the values of the control parameter set {*β*, γ, *α*, *δ*,} in Table 4 determine the nature of the power-law after the exponential growth. On the other hand, the linear regime (for Italy, Spain) near the saturation is determined well by the removal rate set {λ_0_, λ_1_, *κ*_0_, *κ*_1_}. Thus, the present model is consistent with the results presented in Section. 2.

In the next section, we will present another model which is based on delayed differential equations.

## 5 Model based on delayed-differential equations

In this section we consider a class of models based on delay differential equations (DDE), which are different from SEIR model. Here, the equations often look simpler than their SEIR counterparts since delay can be used to account for multiple features without increasing the number of variables. The flip side, however, is that delays can be analytically intractable.

In this section, we focus on one particular delayed model [35], which uses delays to account for the pre-symptomatic period and the infection period. This model has been used to track the evolution of the epidemic, especially in the post-linear regime. It describes a potential new route to the end of the pandemic through a combination of social distancing, sanitization, contact tracing and preventive testing. In the controlled endgame phase of the epidemic, which we call self-burnout, we have a slightly different equation. In this phase, there is extensive enforcement of separation minima (a term we prefer to social distancing as it does not carry connotations of emotional isolation) so the rate of new cases does not depend on the number of healthy and susceptible people at large (i.e. not in quarantine). Rather, we assume that each sick person spreads the disease at a constant rate m_0_. Under these conditions, the dynamic model for the spread of cases (*y*, which is same as *I*(*t*)) is

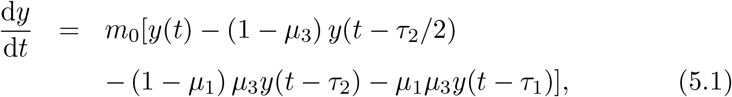

where *μ*_1_,*μ*_2_,*μ*_3_,*τ*_1_,*τ*_2_ are parameter. In our model, the contact tracing manages to capture a fraction 1 − *μ*_3_ of all the sick patients and places them into quarantine.

A solution to the above equation is *y* = *const*. It has been shown in [35] that this solution is stable if and only if

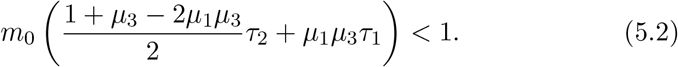

This condition identifies a maximum value of *m*_0_ for which the solution is stable, i.e., the epidemic gets over in time. For the plausible parameter values *τ*_1_ = 7, *τ*^2^ = 3, *μ*_1_ = 1/5 and *μ*_3_ = 1/2, the critical value of *m*_0_ turns out to be 20/53. Here we assume that the test results are instantaneous due to high testing capacity present in the region.

We perform simulation runs of Eq. (5.1) with the above parameter values and *m*_0_ having the values 70%, 80% and 90% of the critical limit. We seed the equation with the linear function *y* = 1000*t* for the first ten days. We find a considerable region thereafter where the case histories show a 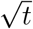 profile before saturating. The errors between the observed data and the best-fit curves are less than 1 percent in each case. This explains why the countries which are achieving saturation are showing a pronounced 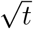 phase after the linear phase.

To further bolster the validity of our model, we consider two countries (South Korea and Austria) which have shown a very good linear region followed by saturation. South Korea showed linear regime from March 28 to April 04 (with 9478 and 10156 cases respectively), after which it enters the burnout phase. Using this as the seeding data and taking the parameter values mentioned above, we find the best fit for the next 20 days for *m*_0_ equal to 77% of the critical. The error between the best-curve and the data is 0.34% and the root mean square error is 0.021. Note that Shayak and Rand [35] has found an *m*_0_ of 75% and not 77 % of critical, because the fit was upto a smaller duration.

Austria showed linear regime from March 28 to April 01 (with 6250 and 10711 cases respectively) before entering self-burnout phase. We find the best fit for *m*_0_ to be 79 % of the critical. The error is 2.6 % and the root mean square error is 0.08. However, the actual data for the 8th to the 15th day appears to be too low—the curve has a convex profile which is probably unrealistic. If we consider the error from the 16th to the 30th day then we find a value of 1.1% only. We present these best-fit curves in Fig. 6. However, in both the cases, a relaxation of lockdown measures initiated in a second wave, which does not fit with the model unless we change the parameters. Hence we have computed the fits only upto the dates when maximal restrictions remained valid. Other regions which are in the self-burnout phase are Vietnam, Australia, New Zealand, and Goa, Kerala and Odisha in India. We have chosen South Korea and Austria since their data shows the smoothest profile on account of high testing capacity.

**Figure 6:**
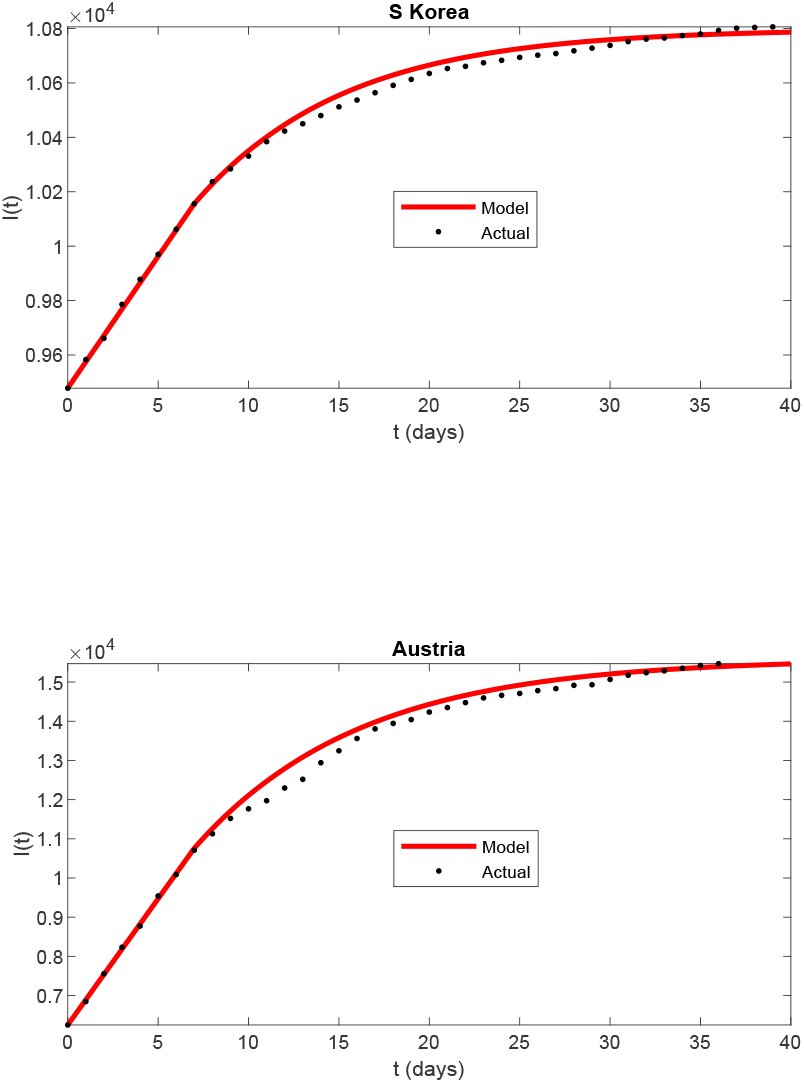
(color online) For the COVID-19 pandemic, *semi-logy* plots of total infected individuals (*I*(*t*)) vs. time (*t*) (black dotted curves) for South Korea and Austria from the available data at *worldOmeter* [40]. The red curves are the best-fit curves obtained using the DDE-based model including self-burnout (see Sec. 5).

Both SEIR and the DDE models describe the evolution of COVID-19 epidemic quite well for many countries. A detailed comparison between the two models will be performed in future. Also, we plan to employ the two models to understand the epidemic evolution for many nations. In the next section, we will sketch a generic scenario for the COVID-19 pandemic evolution that summarizes our previous analysis.

## 6 Effects of lockdown and a generic picture of the pandemic’s evolution

In our current study of 21 countries, we show that the curve depicting the total number of infected individuals (*I*(*t*)) exhibit a power law followed by a linear growth before saturation. We also observe a 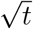 regime between the linear regime and saturation.

In Sec. 4, we observed that for Italy and Spain, the SEIR model satisfactorily captures the variations of *I*(*t*), including the power laws. However, when we turn off the effects of lockdown and social distancing by setting *α* = 0, we observe a steep rise in *I*(*t*) from the exponential regime to saturation. See the dashed curve of Fig. 7 for the *α* = 0 case.

**Figure 7:**
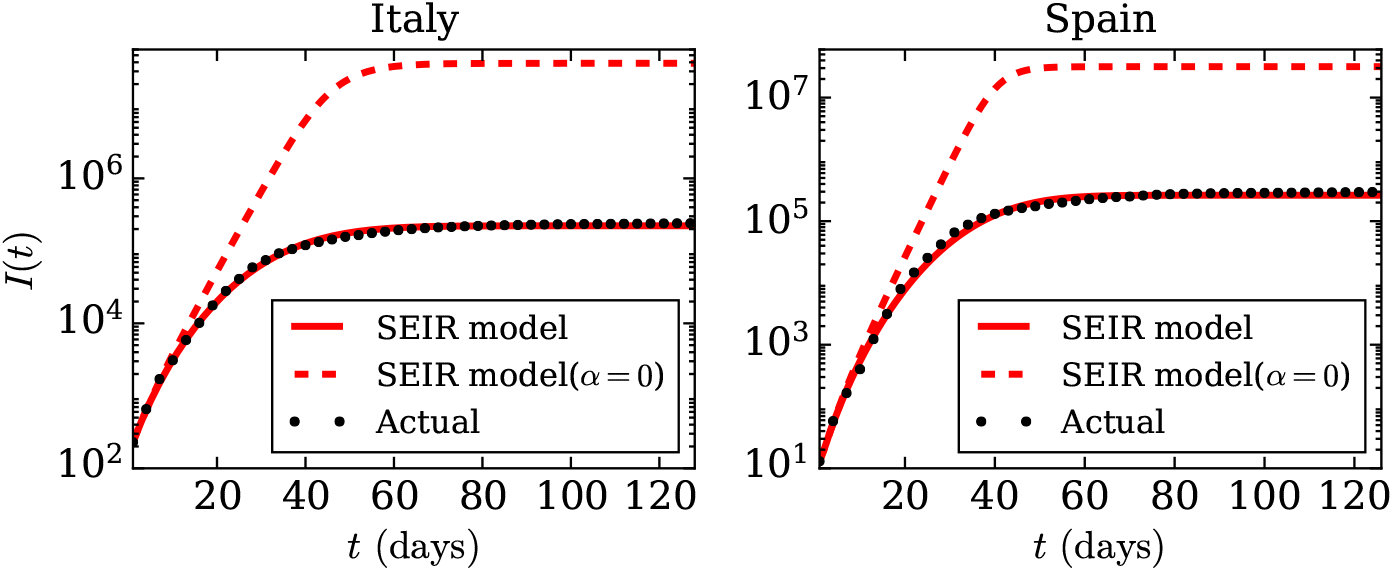
(color online) For the COVID-19 pandemic, *semi-logy* plots of total infected individuals (*I*(*t*)) vs. time (*t*) (black dotted curves) for Italy and Spain from the available data at *worldOmeter* [40] till June 30. The red curves are the best-fit curves using the SEIR model described in Sec. 4. The red dashed curves exhibit the evolution of the total infections without any lockdown or restrictions (*α* = 0).

In Italy, a nationwide lockdown was announced on March 09 [38], while in Spain, a lockdown was imposed on March 14 [39]. In Fig. 7, the separation between the solid and dashed curves occurs around 15th day for both the nations, which is around the time lockdown started for them. Note that *t* = 0 or day-1 corresponds to February 24 for Italy and February 26 for Spain. Thus we show that lockdowns play a vital role in suppressing the peak of the epidemic compared to *α* = 0 case, as well as for introducing power laws in the epidemic curves.

In Fig. 8, we present a schematic diagram that provides a comprehensive picture of our analyses. In the diagram, the solid red curve represents the measured infection counts, whereas the dashed curve depicts *I*(*t*) without intervention. As shown in Fig. 8, the *I*(*t*) curve traces four stages before flattening: *e^βt^*, *t^n^* (where, 2 ≤ *n ≤* 4), *t*, and 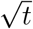, which are labelled as Stage 1, Stage 2, Stage 3, and Stage 4, respectively. Stage 5 represents the final regime where the *I*(*t*) curve is flat. These five regimes are universal, as reported in Sec. 2. In Fig. 8, we also illustrate different regimes of *İ*(*t*) curve using a solid blue line. Note that the stage 3 for which *İ* = *const* (constant daily counts) is the transition zone from convex to concave nature of the epidemic curve. In the final stage, *İ* = 0 (no new cases), correspond to the flattened curve. These unique features of the epidemic curve may be exploited to develop prediction tools for the epidemic evolution.

We conclude in the next section.

**Figure 8:**
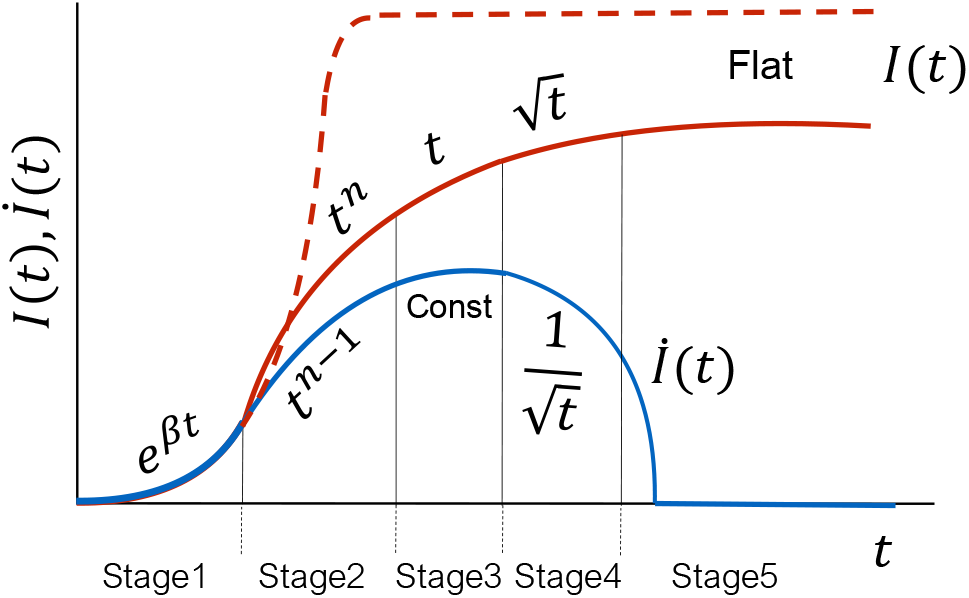
(color online) A schematic plot for time evolution of COVID-19 pandemic in a generic country. The red dashed curve is the path followed by the cumulative infections (*I*(*t*)) without any intervention (lockdown, contact tracing etc.). The solid red curve is the plot for *I*(*t*) with interventions, while the blue curve represents the daily infection cases (*İ*(*t*)). The five stages of the evolution are shown as Stages 1 to 5. The *I*(*t*) curve transitions from convex to concave nature during stage 3. Stage 5 corresponds to the flattened region.

## 7 Discussions and Conclusions

COVID-19 pandemic involves many factors, for example, asymptomatic carriers, lockdown, social distancing, quarantine, etc. Considering these complex issues, we focus on data analysis. In particular, we analyze the real-time infection data of COVID-19 epidemic for 21 nations up to June 30, 2020. Our analysis shows that many nations have flattened the epidemic curve, while some of them (India, Brazil, and Russia) are some distance away from saturation. There is a possibility of a second wave in USA and Israel.

A key feature of our analysis is the emergence of power-law behavior after an exponential growth, which has also been observed by other researchers [42, 17, 23, 5, 26, 19, 36, 2]. We argue that the lockdowns and social distancing may be the possible reasons for the emergence of these power laws. The exponential growth is easily explained using *İ* ∝ *βI* relation, which arises due to the spread by contact. For power-law growth, *I*(*t*) ~ *t^n^*, the above relation is modified to *İ* ~ *I*^1−1/^*^n^*. The suppression of *I*^−1/^*^n^* in *İ* could be attributed to lockdowns and social distancing etc. A careful analysis of the epidemic models should yield this feature. Interestingly, Ebola and MERS also exhibit similar behavior. This generic feature is very useful for the forecast of the epidemic evolution.

Note that the *I*(*t*) curve needs to turn from convex (during the exponential growth) to concave for flattening. Hence, a transition from exponential growth to a power-law growth is expected. The lockdowns and social distancing are likely to make the transition earlier, thus suppressing the exponential growth to some degree. This is also explained in our analysis in Sec. 6. Earlier, Verma et al. [37] had conjectured that the power-law growth might occur due to asymptomatic carriers and/or community spread. This conjecture needs a closer examination.

In this paper, we only studied the infection counts. However, it is evident that during the growth phase, the active cases and death counts, would follow similar pattern as *I*(*t*). The total death count too flattens along with the infection count, but the count of cumulative active cases decreases with time during the saturation.

Prakash et al. [32] studied the phase space portraits, that is, *İ* vs. *I* plots. They observed the phase-space curves to be linear. This is natural for the exponential growth (*İ* ∝ *βI*), as well as for the power-law growth with large exponent *n* because *İ* ~ *I*^1−1/^*^n^*. In another interesting analysis of COVID-19 epidemic, Schuttler et al. [34] and Marsland and Mehta [26] argued that *I*(*t*) or total death count could be modelled using error function. Using this result, we may be able to predict the asymptotic behavior of *I*(*t*) that may yield valuable clues regarding the extent and duration of the epidemic.

Epidemic spread has similarities with rumor spread and the growth of a network [11, 27]. A comparison of the power-law growth in these systems will yield fruitful results for the epidemic forecast.

In summary, COVID-19 epidemic data reveal interesting properties that can be used for its forecast. The 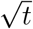 dependence of total infections after the linear regime is a striking feature that might have connections with other aspects of physics and mathematics. We leave these considerations for future studies.

## Data Availability

We used the publicly available data from the website worldOmeter

## Acknowledgments

The authors thank Santosh Ansumali for useful discussions. We also thank the *worldOmeter* for the data that made this work possible. Ali Asad is supported by Indo-French (CEFIPRA) project 6104-1, and Soumyadeep Chatterjee is supported by INSPIRE fellowship (IF180094) of Department of Science & Technology, India.

